# Early pandemic molecular diversity of SARS-CoV-2 in children

**DOI:** 10.1101/2021.02.17.21251960

**Authors:** Ahmed M. Moustafa, William Otto, Xiaowu Gai, Utsav Pandey, Alex Ryutov, Moiz Bootwalla, Dennis T Maglinte, Lishuang Shen, David Ruble, Dejerianne Ostrow, Jeffrey S. Gerber, Jennifer Dien Bard, Rebecca M. Harris, Paul J. Planet

## Abstract

**Background:** In the US, community circulation of the SARS-CoV-2 virus likely began in February 2020 after mostly travel-related cases. Children’s Hospital of Philadelphia began testing on 3/9/2020 for pediatric and adult patients, and for all admitted patients on 4/1/2020, allowing an early glimpse into the local molecular epidemiology of the virus.

**Methods:** We obtained 169 SARS-CoV-2 samples (83 from patients <21 years old) from March through May and produced whole genome sequences. We used genotyping tools to track variants over time and to test for possible genotype associated clinical presentations and outcomes in children.

**Results:** Our analysis uncovered 13 major lineages that changed in relative abundance as cases peaked in mid-April in Philadelphia. We detected at least 6 introductions of distinct viral variants into the population. As a group, children had more diverse virus genotypes than the adults tested. No strong differences in clinical variables were associated with genotypes.

**Conclusions:** Whole genome analysis revealed unexpected diversity, and distinct circulating viral variants within the initial peak of cases in Philadelphia. Most introductions appeared to be local from nearby states. Although limited by sample size, we found no evidence that different genotypes had different clinical impacts in children in this study.

**Summary:** Using sequencing and a novel technique for quantifying SARS-CoV-2 diversity, we investigated 169 SARS-CoV-2 genomes (83 <21 years old). This analysis revealed unexpected diversity especially in children. No clear differences in clinical presentation were associated with the different virus lineages.

## Background

After an initial period in January 2020 when most severe acute respiratory coronavirus 2 (SARS-CoV-2) infections in the US were travel-related, the virus quickly established itself during February with sustained, community spread[1]. Studies tracking the spread of the virus using whole genome phylogenetics suggested multiple introductions during this time period from Europe and Asia [2-7], as well as multiple waves of transmission of distinct variants that differ locally[8].

Understanding genotypic diversity in local molecular epidemiology is critical for tracking spread and new introductions, identifying hotspots, and enhancing contact tracing[2, 4, 5, 7]. However, the biological significance of viral diversity is not known. For instance, it is unclear if lineages differ in virulence or transmissibility[4, 9]. It is also unclear if the immune response will be equally protective against all variants of the virus, highlighting the need to understand SARS-CoV-2 diversity and evolution for vaccine development[2, 10]. Moreover, there is little known about viral diversity across the lifespan, with limited data on SARS-CoV-2 genomic diversity in pediatric populations[11].

The first case of coronavirus disease 2019 (COVID-19) in Philadelphia was reported on March 10, 2020 (https://www.media.pa.gov/pages/health-details.aspx?newsid=734), 14 days after the first non-travel related case was confirmed in California[1] and less than a week after the first cases of community spread in New York State (https://www.governor.ny.gov/news/during-coronavirus-briefing-governor-cuomo-signs-40-million-emergency-management-authorization). On March 9^th^ the infectious disease diagnostic laboratory (IDDL) at Children’s Hospital of Philadelphia (CHOP) became one of the first locations in the region to offer PCR-based testing for SARS-CoV-2, and worked with local authorities to provide testing for both children and adults in the community. On April 1^st^, CHOP instituted universal screening for all admitted children.

To track the molecular epidemiology of the virus locally in Philadelphia, and especially in a pediatric population, we obtained 169 samples from the initial period of testing between 3/19/2020 to 5/4/2020 and performed whole genome sequencing (WGS). Eighty-three samples were from patients less than 21 years old. We used our genotyping tool GNUVID[8] to classify and compare these strains to the growing global database of SARS-CoV-2 sequences at GISAID[12] (Supplementary Table 1)[13]. Here we show that the early pandemic and peak in Philadelphia were characterized by multiple, diverse, circulating viral variants, especially amongst children. We also observed multiple introductions from distinct geographical origins. We report statistics for clinical presentation and outcomes associated with each viral genotype in children.

## Methods

All nasopharyngeal swab samples that had residual volume after initial laboratory processing, from individuals that had positive PCR testing for SARS-CoV-2, were obtained for this study. RNA was extracted from nasopharyngeal swab samples using either the Roche MagNA Pure LC (Roche) or EZ1 virus mini kit (Qiagen) using magnetic bead technology. Whole genome sequencing was done by the Children’s Hospital Los Angeles (CHLA) Center for Personalized Medicine and the Virology Laboratory. Briefly, WGS of extracted viral RNA was performed as previously described using Paragon Genomics CleanPlex SARS-CoV-2 Research and Surveillance NGS Panel[11, 14]. Libraries were quantified using the Agilent High Sensitivity D1000 ScreenTape assay then normalized and pooled on the Biomek i7 liquid handler (Beckman Coulter Life Sciences) to approximately 1nM. The resulting pool was quantified again using the TapeStation High Sensitivity D1000 assay and diluted to a final concentration of 500pM; libraries were denatured and diluted according to Illumina protocols and loaded on the NextSeq 500 at 0.6pM. Paired-end and dual-indexed 2×150bp sequencing was done using NextSeq 500 High Output Kit (300 Cycles).

All SARS-CoV-2 genomes (n=169)[13] were queried against the GNUVID database (version August 17^th^ 2020) that has 32,719 high coverage complete genomes[8, 12]. Each genome was assigned an ST profile and CC. A minimum spanning tree (MST) was then constructed using the goeBURST algorithm[15, 16] to group STs into larger taxonomic units, clonal complexes (CCs), which we define as clusters of >20 STs that are single or double allele variants away from a “founder”[8, 17]. Temporal plots were extracted using a custom script and plotted in GraphPad Prism v7.0a. The genomes were also assigned to a lineage[2] using pangolin (https://github.com/hCoV-2019/pangolin). A custom script was used to check the specific combinations of 9 GISAID genetic markers, and genomes were assigned to the GISAID clades. The genomes were grouped by different age groups and the relative abundance of the STs and the 13 CCs were calculated. To compare the Shannon diversity index between the different groups[18], a t-test was used to determine whether the indices were significantly different[19].

To show the relationship amongst the genomes of the 169 isolates and the global diversity of SARS-CoV-2, a maximum likelihood tree was constructed. Briefly, consensus SARS-CoV-2 sequences for the 169 CHOP isolates were combined with full-length SARS-CoV-2 sequences of 25,807 additional isolates from GISAID[12] that are part of the GNUVID August database release[17] and have an assigned CC and date of isolation (Supplementary Table 1)[13] to generate a multiple sequence alignment using MAFFT’s FFT-NS-2 algorithm[20] (reference MN908947.3[21], options: --add -- keeplength). The 5’ and 3’ untranslated regions were masked in the alignment file using a custom script. A maximum likelihood tree using IQ-TREE 2[22] was then estimated using the HKY model of nucleotide substitution[23], default heuristic search options, and ultrafast bootstrapping with 1000 replicates[24]. The tree was rooted to MN908947.3.

The tree and the six GISAID clades data were visualized in iTOL[25]. The tree and the tip dates were then used in TempEst[26] to estimate the evolutionary rate. Similar procedures were used to construct two trees for both CC4 and CC258 and then estimate the evolutionary rates. Commands used for producing the figures are available in Supplementary Material. Manual review of the electronic health record was performed for all patients who tested positive for SARS-CoV-2 to obtain data on test characteristics, demographic data, exposures, comorbidities, symptomatology, clinical severity, and treatment information and deidentified. Samples were obtained under CHOP IRB protocol 17-014648 as part of routine clinical care, solely for non-research purposes, carrying minimal risk, and were therefore granted a waiver of informed consent. Summary statistics were used to describe demographic and outcome data. Non-parametric methods were used due to our small samples size, and to minimize the effect of outliers on statistical associations. Multivariable logistic regression was used to evaluate the association between viral sequence types and clinical outcomes. All statistics were performed with STATA version 15.0, (Stata Corp., College Station, TX).

## Results

Over the time period of this study, CHOP IDDL performed 4486 tests for SARS-CoV-2 of which 246 (5.48%) were positive. Of the 246 positives in patients <21 years of age, we were able to obtain samples from 71 patients. Of the 71 patients, 15 were admitted, 3 to the intensive care unit (ICU), and 2 needed respiratory support. We also obtained samples from 12 other children and 86 adults tested by the CHOP IDDL for a total of 169 sequences in this study.

Using the GNUVID classifier[8, 12], we genotyped all 169 genomes and assigned a sequence type (ST), which we define as the group of sequences that have exactly the same allelic haplotype. When possible, each ST was then classified into a clonal complex (CC), defined as a group of STs that differ by only one or two alleles from a central “founder” sequence determined by minimum-spanning clustering[8]. Overall, we identified 112 distinct STs in our data, 108 (165 genomes) of which could be assigned to one of 13 CCs when compared to the most recent global GISAID genome database[8, 12, 17]. While 13 STs (56 genomes) had an exact genotype match in the global database, 99 STs (113 genomes) were novel, with previously unobserved alleles that were not due to sequencing ambiguity based on sequence quality. The genomes were widely distributed across the global SARS-CoV-2 phylogeny suggesting multiple introductions (Figure 1A, Supp Fig. 1A). Temporal mapping of the viral CCs by week of isolation showed the persistent predominance of CC258, but also persistence of multiple, diverse haplotypes in the population (Figure 1B).

**Figure 1.**
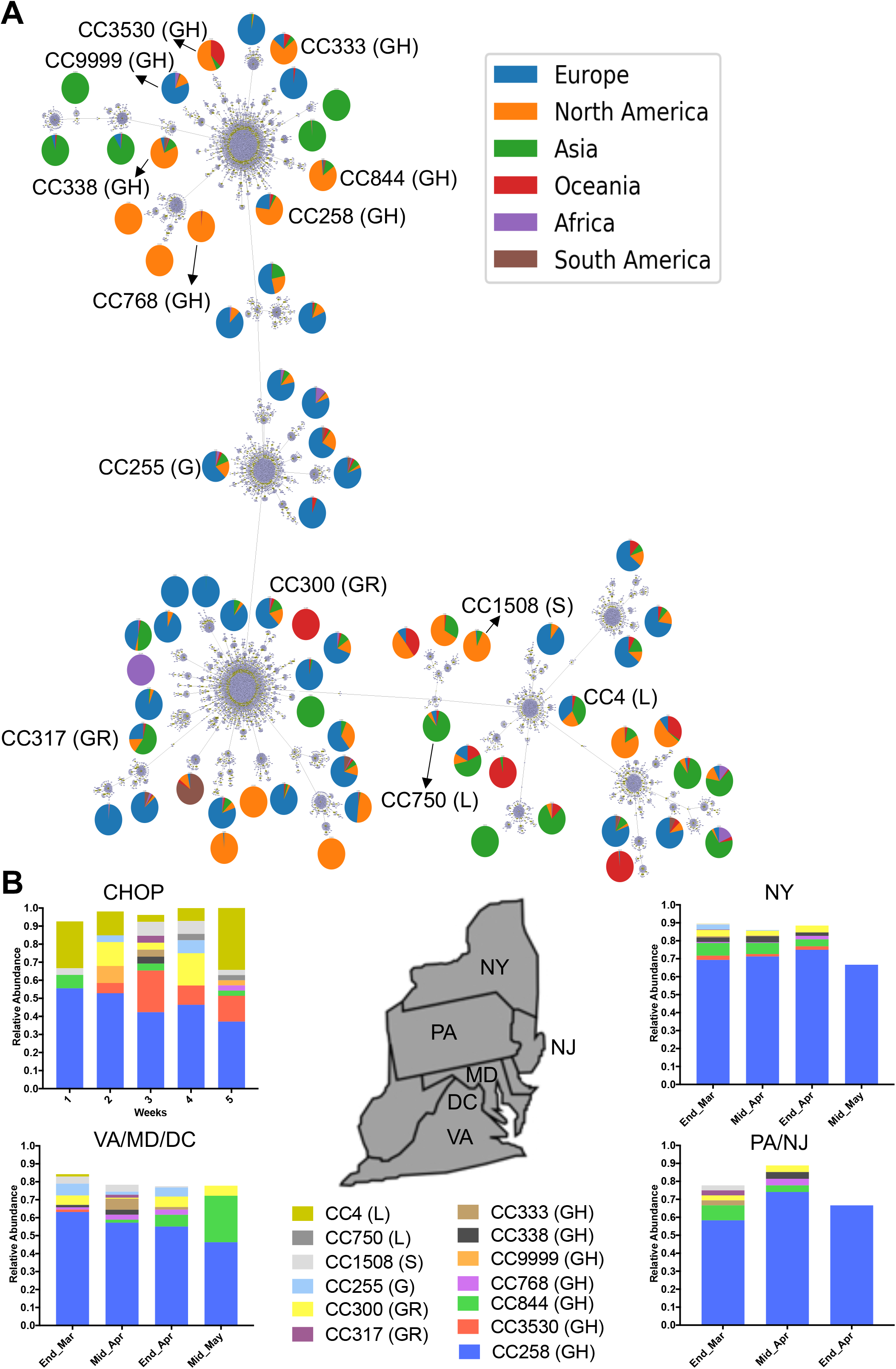
SARS-CoV-2 diversity from testing at our center. **A**. Minimum spanning tree (MST) of 32,719 SARS-CoV-2 genomes showing 17,615 Sequence Types (STs) and 70 clonal complexes (CCs). The MST represents the most recent dataset used in GNUVID as of August 17^th^. The reported 13 CCs at CHOP are in black. The pie charts show the percentage distribution of genomes from the different geographic regions in each CC. **B**. Temporal Plot of 13 circulating CCs representing the 169 genomes in this study and their relative abundance in Pennsylvania (PA) and the neighboring states; New York (NY), New Jersey (NJ), Virginia (VA), Maryland (MD) and District of Columbia (DC). Weeks 1, 2, 3, 4and 5 are from 03/19-03/25, 03/26-04/1, 04/02-04/08, 04/23-04/29 and 04/30-05/04, respectively. The GISAID clades corresponding to the CCs are reported in parentheses.

We estimated the number of putative introductions into our population by comparing our data to high quality sequences from the global GISAID dataset[8, 12, 17], and requiring an identical ST to have been isolated in another geographic location at least 10 days prior to the isolation date in our sample. Using this criterion, we identified 6 independent STs that were likely introductions into our population (Table 1). One of these putatively introduced genotypes, ST6228, had only ever been observed in New York State before, and thus likely represents an introduction from this neighboring state. ST338 and ST258 were also observed in New York State in the 10 days prior to appearing in our population, but they were also widespread internationally during this time period, and therefore could have been introduced from other sources. For ST258, isolates were observed during this time window in 24 countries and 22 States including Pennsylvania and other nearby states such as New Jersey. ST4 and ST1531 were observed closest to Philadelphia in Washington DC and Virginia in the 10 days prior to appearing in our population. The most likely international introduction was ST6134, which was seen previously only in Australia. If we shortened the criterion to isolation 5 days prior, we detected 3 more putative introductions. All 3 of these STs were first observed in New York.

**Table 1:** Introductions to Philadelphia.

| Specimen Date | ST | CC | Days | Countries in last 10 days before appearance | First time Seen\|Date |
| --- | --- | --- | --- | --- | --- |
| 3/19/20 | 4 | 4 | 10 | China Iceland Malaysia Singapore United Kingdom USA (CA, MI, WI) | China/Wuhan\|2019-12-30 |
| 3/24/20 | 258 | 258 | 10 | Australia Austria Canada Chile Colombia Costa Rica Czech Republic Denmark France Germany Greece Iceland Israel Luxembourg Netherlands Portugal Russia Singapore South Korea Sweden Taiwan United Kingdom USA (AZ, CA, CO, CT, FL, GA, IL, IN, ME, MI, MN, NJ, NM, NY, PA, TX, UN, VA, VI, VT, WA, WI) | Singapore\|2020-02-16 |
| 3/30/20 | 1531 | 258 | 10 | Denmark USA (DC, VA, CA) | USA/NY\|2020-03-14 |
| 3/31/20 | 6134 | 258 | 10 | Australia | Australia\|2020-03-19 |
| 3/31/20 | 6228 | 258 | 10 | USA (NY) | USA/NY\|2020-03-21 |
| 4/6/20 | 338 | 338 | 10 | Australia, Colombia, USA (NY, WI, MA, CA, CT, MD, FL) | USA/CA\|2020-02-29 |
| 3/20/20 | 1623 | 258 | 5 | USA (NY) | USA/NY\|2020-03-12 |
| 3/24/20 | 2261 | 258 | 5 | USA (NY) | USA/NY\|2020-03-19 |
| 3/27/20 | 1841 | 3530 | 5 | New Zealand, USA (FL) | USA/NY\|2020-03-18 |

To detect any exportations of viral genotypes, we looked for STs that were seen in our dataset 10 days prior to isolation in another geographic location. Only one possible exportation event was detected of ST13162 to Wisconsin.

It should be noted that our method of detecting introductions relies on robust sampling both in our population and in other locations. The detected number of importations and exportations is likely much higher than the numbers we were able to find here, and estimates may grow as more genome sequences are added from retrospective sampling.

The relative abundance of the 13 CCs found in our dataset was distributed differently between children and adults, with the pediatric population showing considerably more diversity (Shannon Entropy=1.815 vs 1.412, P = 0.0132). CC4, an early lineage originally seen in Wuhan, was more prevalent in pediatric cases (20%) compared to adults (14%). CC258, a lineage that predominated in Europe and New York, was more prevalent in adults (55%) compared to children (40%). A more granular analysis of STs recapitulated the higher diversity of viral types in the pediatric population, but did not achieve statistical significance (Shannon Entropy= 2.624 vs 2.456, P= 0.3557).

One clear difference between our dataset and data from neighboring states over the same time period is the increased diversity of CCs and the presence of the early genotype CC4 (e.g., for NY v. our sample Shannon Entropy=1.69 vs 1.15, P = 4.23E-7). It is unclear whether this reflects specific epidemiology of Philadelphia, our focus on pediatric samples, or other biases in this convenience sample. Interestingly, while there were only 6 STs observed in CC4 (5 STs in children and 2 in adults), there were 57 STs from CC258 (25 STs in children and 38 in adults) demonstrating the much higher diversity of genotypes associated with the CC258 lineage, and potentially the large amount of diversification of this lineage as it peaked to very high numbers in nearby New York City. To address the cause of this diversity, we calculated mutation rates for CC4 and CC258 genomes using our genomes as well as genomes from the GISAID database using TempEst[26] (Supp Fig 1B). The mutation rate for CC4 was 2.2×10^−4^ sites/year while the mutation rate for CC258 was 5.9×10^−4^ sites/year. The rate across all GISAID sequences was 7.1×10^−4^ in line with previous estimates. It is possible that both had a higher mutation rate and a large effective population size through increased transmission contributed to the higher diversity seen in CC258.

To assess the possibility that different genotypes were associated with distinct clinical outcomes and presentations, we collected demographic and clinical information for 71 pediatric viral genomes from patients in the CHOP Care Network. Although limited by the sample size, we were unable to detect any significant differences in specific clinical variables associated with the different genotypes (Tables 2 and 3 and Figure 2). However, exploratory analysis of the data suggested that pediatric patients infected with CC4 lineage virus and early pandemic genotypes (e.g., GISAID lineage L and Pangolin lineage B) may have had increased rates of admission to the hospital (odds ratio, OR 17.2, 95% confidence interval 2.23 to 132.13, P = 0.006) compared to those infected with the CC258 lineage (Supplementary Tables 2, 3 and 4) and lineages considered to be more derived (eg., GISAID lineage GH and Pangolin lineage B.1). In addition, two of the single nucleotide polymorphisms (SNPs) (Table 4 and Supplementary Tables 5 and 6) from more ancestral haplotypes (e.g., C241T, C3037T) were also significantly associated with admission (Supplementary Tables 7, 8, 9 and 10). The D614G (SNP; A23403G) spike protein mutation was associated with less hospital admission, albeit not statistically significant (OR 0.23, 95% CI 0.05-1.13) (Supplementary Table 11), but it was the only SNP tested that was significantly associated with decreased odds of being asymptomatic (OR 0.11, 95% CI 0.01-0.92) (Supplementary Table 12).

**Table 2:** Overall characteristics, grouped by clonal complex (excluding those with single isolate or no clonal complex identified).

|  | Clonal Complex |  |  |  |  |  |  |  |  |
| --- | --- | --- | --- | --- | --- | --- | --- | --- | --- |
|  | Total | CC258 | CC4 | CC3530 | CC300 | CC255 | CC844 | CC1 | CC750 |
|  |  |  |  |  |  |  |  | 508 |  |
|  | 71 | 32 | 10 | 7 | 6 | 4 | 3 | 2 | 2 |
| **Age (years), median** | 10.91 | 11.18 | 7.32 (2.55, | 9.96 (4.73, | 5.37 (.45, | 10.8 (6.6, | 8.84 | 16.5 | 8.5 (7.6, |
| **(IQR)** | (5.6, 17.0) | (7.1, 17.2) | 14.75) | 18.11) | 13.19) | 12.8) | (8.84, 8.84) | (15.8 , 19) | 9.3) |
| **Age Group** |  |  |  |  |  |  |  |  |  |
| **0-12 months** | 6 (8%) | 1 (3%) | 1 (10%) | 1 (14%) | 3 (50%) | 0 (0%) | 0 (0%) | 0 | 0 (0%) |
|  |  |  |  |  |  |  |  | (0%) |  |
| **1-5 years** | 12 (17%) | 6 (19%) | 4 (40%) | 1 (14%) | 0 (0%) | 1 (25%) | 0 (0%) | 0 | 0 (0%) |
|  |  |  |  |  |  |  |  | (0%) |  |
| **6-11 years** | 20 (28%) | 10 (31%) | 1 (10%) | 2 (29%) | 1 (17%) | 2 (50%) | 1 (100%) | 0 | 2 |
|  |  |  |  |  |  |  |  | (0%) | (100%) |
| **12-18 years** | 24 (34%) | 12 (38%) | 4 (40%) | 1 (14%) | 2 (33%) | 1 (25%) | 0 (0%) | 2 | 0 (0%) |
|  |  |  |  |  |  |  |  | (67 %) |  |
| **18-21 years** | 9 (13%) | 3 (9%) | 0 (0%) | 2 (29%) | 0 (0%) | 0 (0%) | 0 (0%) | 1 | 0 (0%) |
|  |  |  |  |  |  |  |  | (33 %) |  |
| **Male sex** | 32 (45%) | 13 (41%) | 4 (40%) | 5 (71%) | 3 (50%) | 3 (75%) | 2 (67%) | 0 | 1 (50%) |
|  |  |  |  |  |  |  |  | (0%) |  |
| **Race/Ethnicity** |  |  |  |  |  |  |  |  |  |
| **Non-Hispanic White** | 19 (27%) | 8 (25%) | 5 (50%) | 0 (0%) | 2 (33%) | 0 (0%) | 1 (100%) | 0 | 1 (50%) |
|  |  |  |  |  |  |  |  | (0%) |  |
| **Non-Hispanic Black** | 38 (54%) | 18 (56%) | 3 (30%) | 7 (100%) | 2 (33%) | 3 (75%) | 0 (0%) | 2 | 1 (50%) |
|  |  |  |  |  |  |  |  | (67 %) |  |
| **Hispanic or Latino** | 7 (10%) | 3 (9%) | 1 (10%) | 0 (0%) | 1 (17%) | 1 (25%) | 0 (0%) | 0 | 0 (0%) |
|  |  |  |  |  |  |  |  | (0%) |  |
| **Multi-racial** | 2 (3%) | 1 (3%) | 1 (10%) | 0 (0%) | 0 (0%) | 0 (0%) | 0 (0%) | 0 | 0 (0%) |
|  |  |  |  |  |  |  |  | (0%) |  |
| **Hawaiian or Pacific**<br>**Islander** | 1 (1%) | 0 (0%) | 0 (0%) | 0 (0%) | 0 (0%) | 0 (0%) | 0 (0%) | 1<br>(33<br>%) | 0 (0%) |
| **Other Race or**<br>**Unknown** | 4 (6%) | 2 (6%) | 0 (0%) | 0 (0%) | 1 (17%) | 0 (0%) | 0 (0%) | 0<br>(0%) | 0 (0%) |
| **Insurance status** |  |  |  |  |  |  |  |  |  |
| **Commercial**<br>**Insurance** | 26 (37%) | 13 (41%) | 4 (40%) | 3 (43%) | 2 (33%) | 0 (0%) | 0 (0%) | 0<br>(0%) | 0 (0%) |
| **Government or**<br>**Public Insurance** | 40 (56%) | 18 (56%) | 4 (40%) | 4 (57%) | 4 (67%) | 4 (100%) | 1 (100%) | 2<br>(67<br>%) | 1 (50%) |
| **Self-pay** | 1 (1%) | 0 (0%) | 0 (0%) | 0 (0%) | 0 (0%) | 0 (0%) | 0 (0%) | 0<br>(0%) | 1 (50%) |
| **Other or Unknown** | 4 (6%) | 1 (3%) | 2 (20%) | 0 (0%) | 0 (0%) | 0 (0%) | 0 (0%) | 1<br>(33<br>%) | 0 (0%) |
| **Previously Healthy** | 23 (32%) | 12 (38%) | 3 (30%) | 2 (29%) | 2 (33%) | 1 (25%) | 0 (0%) | 1<br>(33<br>%) | 1 (50%) |
| **Admitted** | 15 (21%) | 3 (9%) | 5 (50%) | 3 (43%) | 1 (17%) | 1 (25%) | 1 (33%) | 0<br>(0%) | 1 (50%) |
| **ICU admission** | 3 (4%) | 0 (0%) | 0 (0%) | 0 (0%) | 1 (17%) | 1 (25%) | 0 (0%) | 0<br>(0%) | 1 (50%) |
| **Need for respiratory**<br>**support** | 2 (3%) | 0 (0%) | 1 (10%) | 0 (0%) | 1 (17%) | 0 (0%) | 0 (0%) | 0<br>(0%) | 0 (0%) |
| **Clinical Severity** |  |  |  |  |  |  |  |  |  |
| **Asymptomatic** | 7 (10%) | 2 (6%) | 3 (30%) | 1 (14%) | 1 (17%) | 0 (0%) | 0 (0%) | 0<br>(0%) | 0 (0%) |
| **Mild** | 60 (86%) | 29 (94%) | 6 (60%) | 6 (86%) | 4 (67%) | 4 (100%) | 3 (100%) | 2<br>(100<br>%) | 1 (50%) |
| **Severe** | 3 (4%) | 0 (0%) | 1 (10%) | 0 (0%) | 1 (17%) | 0 (0%) | 0 (0%) | 0<br>(0%) | 1 (50%) |

**Table 3:**
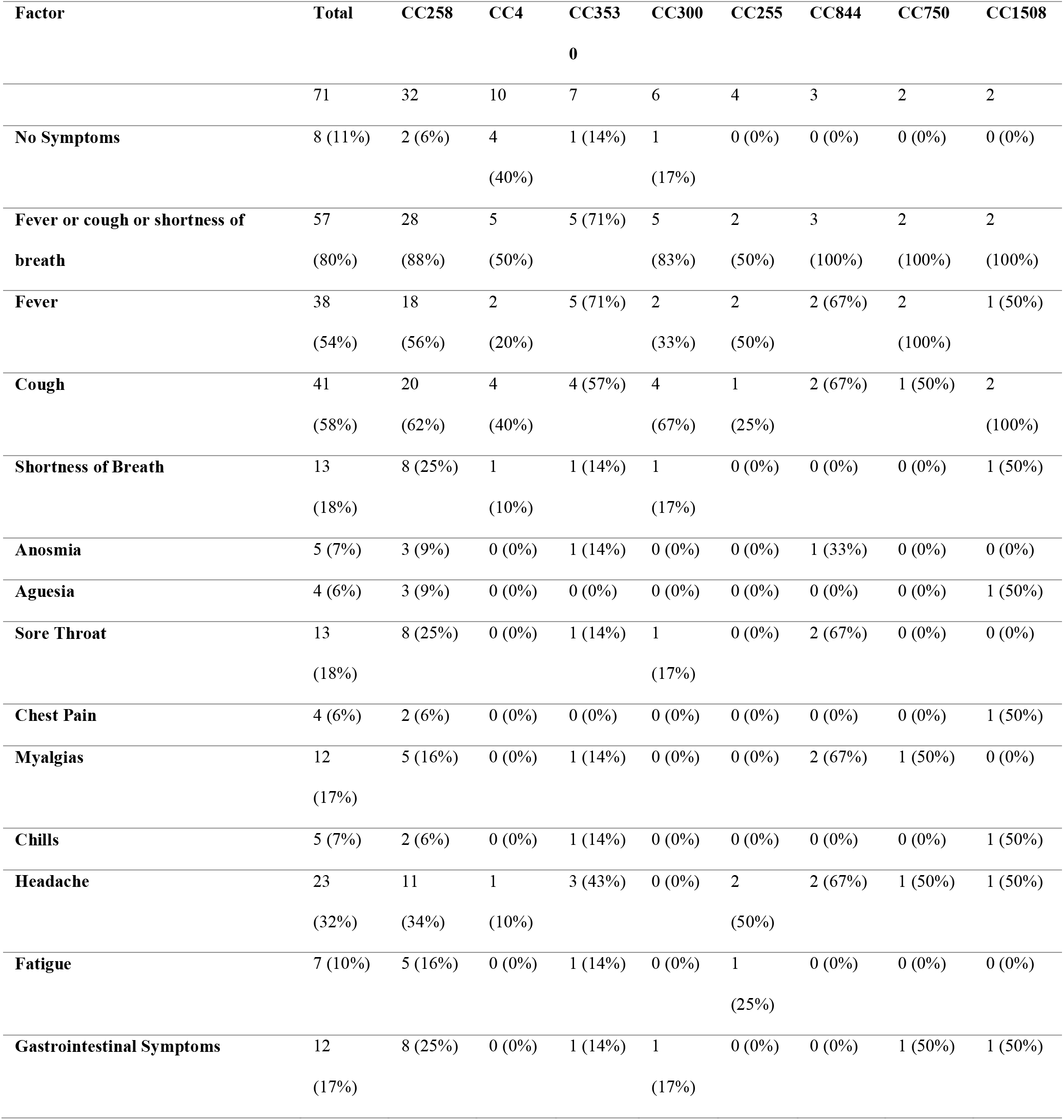
Symptoms, grouped by clonal complex (excluding those with single isolate or no clonal complex identified).

**Table 4:**
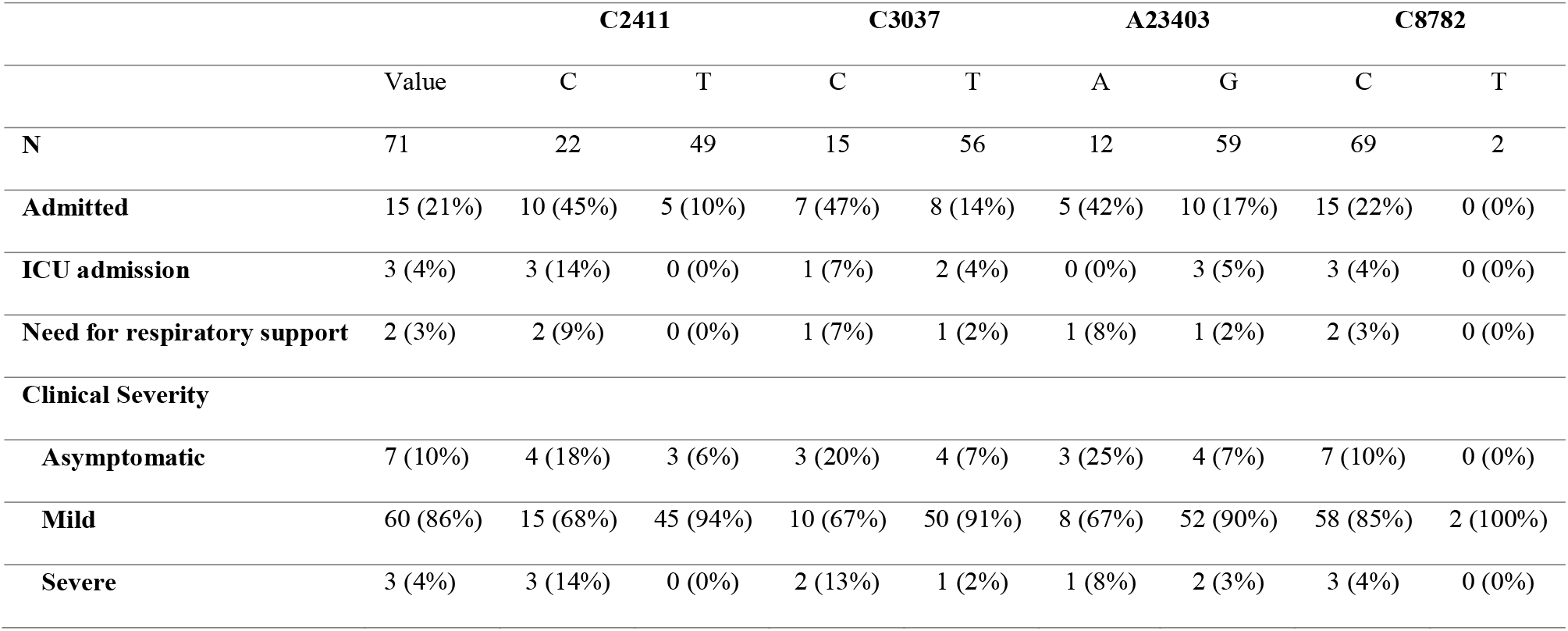
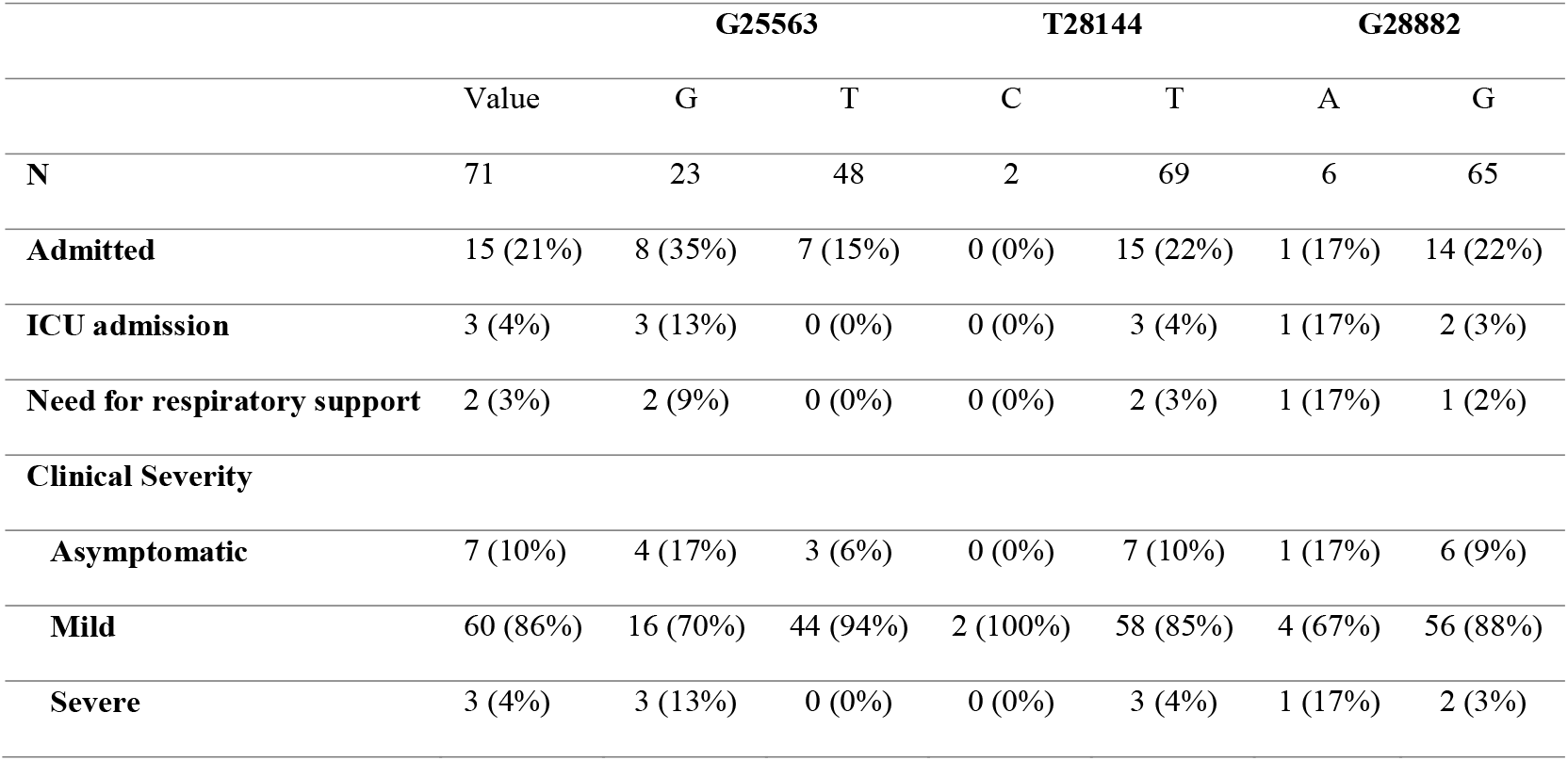
Outcomes, grouped by SNP (excluding those with single type)

**Figure 2.**
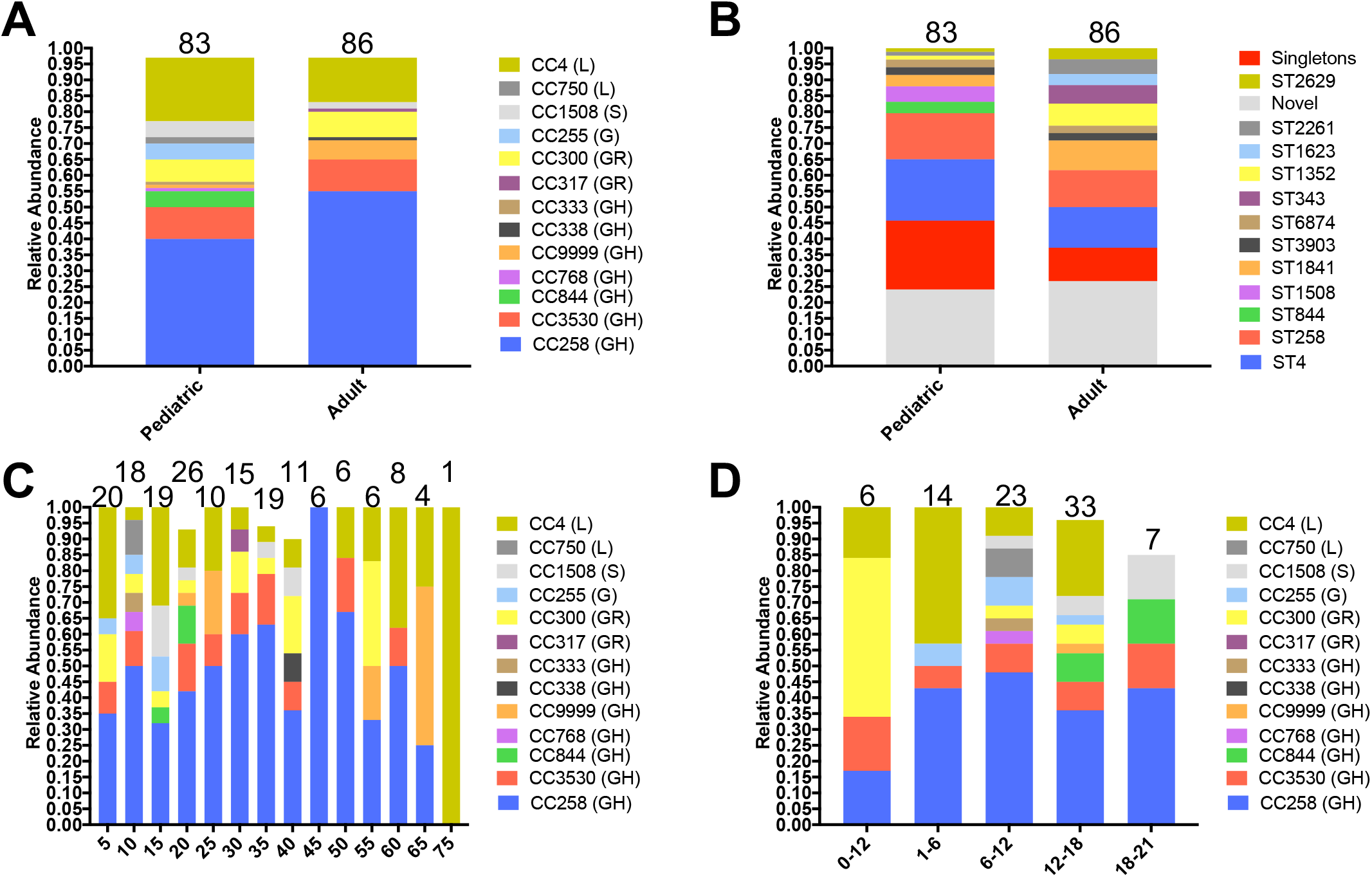
SARS-CoV-2 diversity across different age groups in our sample. **A**. Relative abundance of circulating CCs between pediatrics (≤ 21 years old) and adults. **B**. Relative abundance of circulating STs between children (≤ 21 years old) and adults. **C**. Relative abundance of circulating CCs in 5-year age groups. **D**. Relative abundance of circulating CCs in childhood age ranges (≤ 21 years old). Relative abundance is the ratio of the number of genomes belonging a certain CC (lineage) divided by the total number of genomes in a certain time window. The numbers on the bars represent the total number of genomes in each group.

## Discussion

We have shown that the early pandemic in Philadelphia was diverse and dynamic, with multiple likely introductions, most probably from local spread of the virus from neighboring states. Although CC258, the clonal complex thought to have been introduced from Europe that dominated in New York[4, 8], also predominated in our sample across the early pandemic, other CCs were robustly present. For instance, CC4, one of the earliest genotypes seen in Wuhan, persisted throughout the study period demonstrating sustained spread in the community. Other CCs (e.g., CC3530, CC300, CC1508) were also seen persistently in this sample implying sustained community spread. This finding suggests that there was enough viral diversity early in the pandemic that contact tracing may have been significantly enhanced by whole genome (or targeted SNP detection) comparisons.

It is important to note that most of the putative introductions into our population could be traced to nearby states surrounding the Philadelphia area, and only one putative international introduction was detected. This may reflect international travel restrictions in place at this time, but it also suggests that most spread was local, and that there were missed opportunities to limit these events particularly in travel to and from New York. It is important to note that as the database of SARS-CoV-2 genomes grows and more genome sequences are available from the Philadelphia area, we may find new evidence for introductions or importations, which likely far outnumber those detected in our analysis.

Although the viral genotypes in our sample differed at several putatively key amino acid locations, we did not detect any stark differences in clinical presentation or outcome in children (Tables 2 and 3). Previous studies have shown that different nucleotide variants or deletions may be associated with higher or lower severity [27, 28]. However, the small sample size and higher than expected viral diversity might have led to an inability to discriminate smaller effect sizes. It should also be noted that the retrospective nature of this study, incomplete sampling, and inconsistent capture of symptoms and severity, could have biased these data. Nonetheless, it is still possible that genetic differences between viral lineages may have an impact on virulence or clinical outcome, and our observed differences in admission rates raises the possibility that larger studies may uncover differences in the future. Notably, another recent pediatric study of 141 SARS-CoV-2 in California, which assessed clinical characteristics of 88 patients, demonstrated a possible association between a specific genotype and disease severity[11].

It is also possible that genetic variants may have differential transmission abilities, which could not have been detected directly using our data. However, it is worth noting that the genotypes (CC4, CC750 and CC1508) that have the ancestral alanine residue at position 614 in the spike protein persisted and spread throughout the study period, suggesting that the derived allelic form (A23403G; D614G) that has been proposed to be more transmissible[29] and is predominantly represented by CC258 in our analysis, did not completely dominate the ancestral form over this amount of time. Here we also showed much higher diversity in the CC258 lineage and a higher estimated mutation for this CC in general. It is possible that this diversity is driven by higher transmissibility and a large effective population size.

Overall, our findings suggest that whole genome sequencing and genotyping of circulating clones could be used to track viral spread and identify opportunities for intervention to stop spread from specific hotspots. The relationship between viral genotype, rate of transmission, and clinical presentation and outcomes deserves further exploration with increased sample size.

## Data Availability

The 169 genomes from our dataset will be available from the corresponding author and available online for download through a permanent Zenodo DOI[13]. Other de-identified clinical data used in the manuscript are available upon request from the corresponding author. The GNUVID compressed database and GNUVID source code can be found in its most up-to-date version here, https://github.com/ahmedmagds/GNUVID, under the GNU General Public License.

DOI:10.5281/zenodo.4282048

## List of abbreviations

GNUVID: Gene Novelty Unit-based Virus Identification
ST: Sequence Type
CC: Clonal Complex
SARS-CoV-2: Severe Acute Respiratory Syndrome Corona Virus 2
COVID-19: Corona Virus Disease 2019
wgMLST: whole genome Multilocus Sequence Typing

## Authors’ contributions

AMM & PJP designed and conceptualized the study. AMM contributed to the data collection, data analysis, coding, data interpretation, figures, literature review and tables. WO contributed to the data collection, data analysis, data interpretation, tables, and writing. XG, UP, AR, MB, DTM, LS, DR and DO contributed to the data collection, data analysis, and data interpretation. JDB, JSG, RMH and PJP supervised the study and contributed to the data collection, data analysis, data interpretation, and literature review. AMM and PJP wrote the first draft of the manuscript. All authors reviewed and approved the final manuscript.

## Acknowledgements

We would like to acknowledge the staff members of the infectious disease diagnostic laboratory (IDDL) at the Children’s Hospital of Philadelphia (CHOP) and the Virology laboratory and Center for Personalized Medicine at Children’s Hospital Los Angeles for processing and sequencing the isolates. We would also like to thank the Global Initiative on Sharing All Influenza Data (GISAID) and thousands of contributing laboratories for making the genomes publicly available. A full acknowledgements table is available through GNUVID[17].

