## Supplementary Methods, Tables and Figures for "Early pandemic molecular diversity of SARS-CoV-2 in children"

**Supplementary Material**

Commands for Figure 1	2

Figure 1A	2

Figure 1B	2

Commands for Figure 2	2

Commands for Supplementary Figure 1	2

References	3

Supplementary Figure 1	4

Supplementary Table 2	5

Supplementary Table 3	6

Supplementary Table 4	7

Supplementary Table 5	8

Supplementary Table 6	11

Supplementary Table 7	13

Supplementary Table 8	14

Supplementary Table 9	15

Supplementary Table 10	16

Supplementary Table 11	17

Supplementary Table 12	18

### Commands for Figure 1

#### Figure 1A

The 169 genomes from this study were queried against the GNUVID database (version August 17th 2020) that has 32,719 high coverage complete genomes^1,2^. Each genome was assigned an ST profile and CC. The 13 CCs that were assigned to the 169 genomes were mapped on the minimum spanning tree of the 32,719 high coverage complete SARS-CoV-2 genomes.

*GNUVID.py -d GNUVID_08172020_comp_db.txt MN908947.3_cds.fna WG folder_169_genomes/*

#### Figure 1B

The records for all isolates from NY, VA, DC, MD, NJ and PA between March 16^th^ and May 15^th^ were extracted from the GNUVID August report^2^. The following command was used with each state and with the 169 genomes. The temporal stacked bar plots were then produced in GraphPad Prism v7.0a.

*extract_US_genomes.py MD NJ PA VA DC GNUVID_08172020_DB_isolates_report.txt*

*stacked_area_graph.py -l CHOP_time_order.txt CHOP_GNUVID_time_reports/*

*stacked_area_graph.py -l VA_MD_DC_order.txt VA_MD_DC_GNUVID_time_reports/*

*stacked_area_graph.py -l NY_order.txt NY_GNUVID_time_reports /*

*stacked_area_graph.py -l PA_NJ_order.txt PA_NJ_GNUVID_time_reports /*

### Commands for Figure 2

The genomes were grouped by different age groups and the relative abundance of the STs and the 13 CCs were calculated. The stacked bar plots were then produced in GraphPad Prism v7.0a. The following command was used for each age group.

*Age_figures.py Age_1_year.txt Age_CCs.txt*

### Commands for Supplementary Figure 1

A multiple sequence alignment was constructed using MAFFT’s FFT-NS-2 algorithm21 (reference MN908947.322, options: --add --keeplength). The specific combinations of 9 GISAID genetic markers were then checked, and genomes were assigned to the GISAID clades. The 5’ and 3’ untranslated regions were masked in the alignment file. A maximum likelihood tree was constructed, including the 169 CHOP isolates and 25,807 additional isolates from GISAID that are part of the GNUVID August database release and have an assigned CC and date of isolation (Supplementary Table 1). Two other maximum likelihood trees for CC4 and CC258 were constructed. The trees and the tip dates were then used in TempEst to estimate the evolutionary rate.

*mafft --thread 2 --add 169_genomes.fna --keeplength MN908947.3.fna > 169_genomes_aligned.fna*

*SNPs_finder_CHLA_CHOP.py 169_genomes_aligned.fna*

*mask_regions_alignment.py 169_genomes_aligned.fna 169_genomes_aligned_maksed.fna*

*iqtree -st DNA -m HKY -B 1000 --nmax 100 -T 32 -s Multifasta_25977_aligned_169_added_masked.fna*

*iqtree -st DNA -m HKY -B 1000 --nmax 100 -s Multifasta_1148_CC4_aligned_masked.fna*

*iqtree -st DNA -m HKY -B 1000 --nmax 100 -s Multifasta_5803_CC258_aligned_masked.fna*

Supplementary Figure 1**. The study isolates in the context of the SARS-CoV-2 global diversity.**

**A.** Maximum likelihood phylogeny of 25,977 SARS-CoV-2 genomes. These genomes include 169 genomes from this study and 25,808 global high-quality SARS-CoV-2 sequences downloaded from the GISAID database (http://gisaid.org) on August 17^th^ 2020 (Supplementary Table 3). The tree is rooted on the reference sequence MN908947.3. First Ring and Second Ring from inside to outside represent the six GISAID clades and the 169 study isolates, respectively. The tree was visualized in iTOL. The raw tree is available from the authors. **B.** Estimated evolutionary rate for 25,977 SARS-CoV-2 genomes using TempEst.

Supplementary Table 2**. Comparison for hospital admission of clonal complex, 258 versus 4.**

|  | Odds Ratio | 95% Confidence Interval | P-value |
| --- | --- | --- | --- |
| Clonal Complex |  |  |  |
| CC256 | -ref- |  |  |
| CC4 | 13.14 | 1.61-107.62 | 0.016 |
| Age in years | 0.93 | 0.78-1.11 | 0.444 |
| Sex |  |  |  |
| Female | -ref- |  |  |
| Male | 0.89 | 0.10-8.14 | 0.918 |
| Race/Ethnicity |  |  |  |
| Non-Hispanic White | -ref- |  |  |
| Non-Hispanic Black | 1.15 | 0.09-14.50 | 0.915 |
| Other Race or Ethnicity | 0.74 | 0.02-24.72 | 0.868 |
| Insurance |  |  |  |
| Commercial Insurance | -ref- |  |  |
| Government or Public Insurance | 0.96 | 0.09-10.28 | 0.973 |

Model: Clonal complex, age in years at time of testing, sex, race/ethnicity, and insurance payer

Supplementary Table 3**. Comparison for Hospital admission of Pangolin lineage, B.1 versus B.**

|  | Odds Ratio | 95% Confidence Interval | P-value |
| --- | --- | --- | --- |
| Lineage |  |  |  |
| B.1 | -ref- |  |  |
| B | 17.15 | 2.23-132.13 | 0.006 |
| Age in years | 0.98 | 0.84-1.15 | 0.804 |
| Sex |  |  |  |
| Female | -ref- |  |  |
| Male | 1.52 | 0.23-9.95 | 0.660 |
| Race/Ethnicity |  |  |  |
| Non-Hispanic White | -ref- |  |  |
| Non-Hispanic Black | 1.12 | 0.12-10.36 | 0.923 |
| Other Race or Ethnicity | 0.47 | 0.02-9.81 | 0.629 |
| Insurance |  |  |  |
| Commercial Insurance | -ref- |  |  |
| Government or Public Insurance | 1.58 | 0.21-11.72 | 0.656 |

Model: Viral lineage, age in years at time of testing, sex, race/ethnicity, and insurance payer

Supplementary Table 4**. Comparison for Hospital admission of GISAID clade, GH versus L.**

|  | Odds Ratio | 95% Confidence Interval | P-value |
| --- | --- | --- | --- |
| GISAID Clade |  |  |  |
| GH | -ref- |  |  |
| L | 6.64 | 1.10-40.04 | 0.039 |
| Age in years | 0.97 | 0.85-1.11 | 0.680 |
| Sex |  |  |  |
| Female | -ref- |  |  |
| Male | 1.82 | 0.39-8.55 | 0.449 |
| Race/Ethnicity |  |  |  |
| Non-Hispanic White | -ref- |  |  |
| Non-Hispanic Black | 0.84 | 0.13-5.54 | 0.859 |
| Other Race or Ethnicity | 0.21 | 0.01-3.19 | 0.260 |
| Insurance |  |  |  |
| Commercial Insurance | -ref- |  |  |
| Government or Public Insurance | 2.90 | 0.51-16.51 | 0.230 |

Model: GISAID clade, age in years at time of testing, sex, race/ethnicity, and insurance payer

Supplementary Table 5**: Overall characteristics, grouped by SNP (excluding those with single type)**

|  |  | C241 | | C3037 | | A23403 | | C8782 | |
| --- | --- | --- | --- | --- | --- | --- | --- | --- | --- |
|  | Total | C | T | C | T | A | G | C | T |
| N | 71 | 22 | 49 | 15 | 56 | 12 | 59 | 69 | 2 |
| Age (y), median (IQR) | 10.9 (5.6, 17.0) | 9.3 (3.1, 14.8) | 11.5 (7.4, 17.2) | 9.0 (2.6, 14.8) | 11.4 (7.2, 17.2) | 10.7 (2.6, 15.8) | 10.9 (7.1, 17.2) | 10.7 (5.6, 17.0) | 16.2 (12.3, 20.0) |
| Age Group |  |  |  |  |  |  |  |  |  |
| 0-12 months | 6 (8%) | 2 (9%) | 4 (8%) | 1 (7%) | 5 (9%) | 1 (8%) | 5 (8%) | 6 (9%) | 0 (0%) |
| 1-5 years | 12 (17%) | 6 (27%) | 6 (12%) | 5 (33%) | 7 (12%) | 4 (33%) | 8 (14%) | 12 (17%) | 0 (0%) |
| 6-11 years | 20 (28%) | 5 (23%) | 15 (31%) | 3 (20%) | 17 (30%) | 1 (8%) | 19 (32%) | 20 (29%) | 0 (0%) |
| 12-18 years | 24 (34%) | 7 (32%) | 17 (35%) | 5 (33%) | 19 (34%) | 5 (42%) | 19 (32%) | 23 (33%) | 1 (50%) |
| 18-21 years | 9 (13%) | 2 (9%) | 7 (14%) | 1 (7%) | 8 (14%) | 1 (8%) | 8 (14%) | 8 (12%) | 1 (50%) |
| Male sex | 32 (45%) | 9 (41%) | 23 (47%) | 6 (40%) | 26 (46%) | 4 (33%) | 28 (47%) | 32 (46%) | 0 (0%) |
| Race/Ethnicity |  |  |  |  |  |  |  |  |  |
| Non-Hispanic White | 19 (27%) | 7 (32%) | 12 (24%) | 7 (47%) | 12 (21%) | 6 (50%) | 13 (22%) | 18 (26%) | 1 (50%) |
| Non-Hispanic Black | 38 (54%) | 11 (50%) | 27 (55%) | 6 (40%) | 32 (57%) | 4 (33%) | 34 (58%) | 37 (54%) | 1 (50%) |
| Hispanic or Latino | 7 (10%) | 2 (9%) | 5 (10%) | 1 (7%) | 6 (11%) | 1 (8%) | 6 (10%) | 7 (10%) | 0 (0%) |
| Multi-racial | 2 (3%) | 1 (5%) | 1 (2%) | 1 (7%) | 1 (2%) | 1 (8%) | 1 (2%) | 2 (3%) | 0 (0%) |
| Hawaiian or Pacific Islander | 1 (1%) | 0 (0%) | 1 (2%) | 0 (0%) | 1 (2%) | 0 (0%) | 1 (2%) | 1 (1%) | 0 (0%) |
| Other Race or Unknown | 4 (6%) | 1 (5%) | 3 (6%) | 0 (0%) | 4 (7%) | 0 (0%) | 4 (7%) | 4 (6%) | 0 (0%) |
| Insurance status |  |  |  |  |  |  |  |  |  |
| Commercial | 26 (37%) | 5 (23%) | 21 (43%) | 5 (33%) | 21 (38%) | 5 (42%) | 21 (36%) | 25 (36%) | 1 (50%) |
| Government | 40 (56%) | 14 (64%) | 26 (53%) | 7 (47%) | 33 (59%) | 5 (42%) | 35 (59%) | 39 (57%) | 1 (50%) |
| Self-pay | 1 (1%) | 1 (5%) | 0 (0%) | 1 (7%) | 0 (0%) | 0 (0%) | 1 (2%) | 1 (1%) | 0 (0%) |
| Other or Unknown | 4 (6%) | 2 (9%) | 2 (4%) | 2 (13%) | 2 (4%) | 2 (17%) | 2 (3%) | 4 (6%) | 0 (0%) |
| Previously Healthy | 23 (32%) | 6 (27%) | 17 (35%) | 4 (27%) | 19 (34%) | 3 (25%) | 20 (34%) | 23 (33%) | 0 (0%) |
| Admitted | 15 (21%) | 10 (45%) | 5 (10%) | 7 (47%) | 8 (14%) | 5 (42%) | 10 (17%) | 15 (22%) | 0 (0%) |
| ICU admission | 3 (4%) | 3 (14%) | 0 (0%) | 1 (7%) | 2 (4%) | 0 (0%) | 3 (5%) | 3 (4%) | 0 (0%) |
| Need for respiratory support | 2 (3%) | 2 (9%) | 0 (0%) | 1 (7%) | 1 (2%) | 1 (8%) | 1 (2%) | 2 (3%) | 0 (0%) |
| Clinical Severity |  |  |  |  |  |  |  |  |  |
| Asymptomatic | 7 (10%) | 4 (18%) | 3 (6%) | 3 (20%) | 4 (7%) | 3 (25%) | 4 (7%) | 7 (10%) | 0 (0%) |
| Mild | 60 (86%) | 15 (68%) | 45 (94%) | 10 (67%) | 50 (91%) | 8 (67%) | 52 (90%) | 58 (85%) | 2 (100%) |
| Severe | 3 (4%) | 3 (14%) | 0 (0%) | 2 (13%) | 1 (2%) | 1 (8%) | 2 (3%) | 3 (4%) | 0 (0%) |

**Supplementary Table 5: Overall characteristics, grouped by SNP (excluding those with single type)**

|  | T28144 | | G288882 | | G25563 | |
| --- | --- | --- | --- | --- | --- | --- |
| Factor | C | T | A | G | G | T |
| N | 2 | 69 | 6 | 65 | 23 | 48 |
| Age (y), median (IQR) | 16.2 (12.3, 20.00) | 10.7 (5.6, 17.0) | 5.37 (.45, 13.19) | 11.3 (7.1, 17.1) | 10.0 (2.6, 14.2) | 11.9 (7.5, 17.3) |
| Age Group |  |  |  |  |  |  |
| 0-12 months | 0 (0%) | 6 (9%) | 3 (50%) | 3 (5%) | 4 (17%) | 2 (4%) |
| 1-5 years | 0 (0%) | 12 (17%) | 0 (0%) | 12 (18%) | 5 (22%) | 7 (15%) |
| 6-11 years | 0 (0%) | 20 (29%) | 1 (17%) | 19 (29%) | 5 (22%) | 15 (31%) |
| 12-18 years | 1 (50%) | 23 (33%) | 2 (33%) | 22 (34%) | 8 (35%) | 16 (33%) |
| 18-21 years | 1 (50%) | 8 (12%) | 0 (0%) | 9 (14%) | 1 (4%) | 8 (17%) |
| Male sex | 0 (0%) | 32 (46%) | 3 (50%) | 29 (45%) | 10 (43%) | 22 (46%) |
| Race/Ethnicity |  |  |  |  |  |  |
| Non-Hispanic White | 1 (50%) | 18 (26%) | 2 (33%) | 17 (26%) | 9 (39%) | 10 (21%) |
| Non-Hispanic Black | 1 (50%) | 37 (54%) | 2 (33%) | 36 (55%) | 9 (39%) | 29 (60%) |
| Hispanic or Latino | 0 (0%) | 7 (10%) | 1 (17%) | 6 (9%) | 3 (13%) | 4 (8%) |
| Multi-racial | 0 (0%) | 2 (3%) | 0 (0%) | 2 (3%) | 1 (4%) | 1 (2%) |
| Hawaiian or Pacific Islander | 0 (0%) | 1 (1%) | 0 (0%) | 1 (2%) | 0 (0%) | 1 (2%) |
| Other Race or Unknown | 0 (0%) | 4 (6%) | 1 (17%) | 3 (5%) | 1 (4%) | 3 (6%) |
| Insurance status |  |  |  |  |  |  |
| Commercial | 1 (50%) | 25 (36%) | 2 (33%) | 24 (37%) | 7 (30%) | 19 (40%) |
| Government | 1 (50%) | 39 (57%) | 4 (67%) | 36 (55%) | 14 (61%) | 26 (54%) |
| Self-pay | 0 (0%) | 1 (1%) | 0 (0%) | 1 (2%) | 0 (0%) | 1 (2%) |
| Other or Unknown | 0 (0%) | 4 (6%) | 0 (0%) | 4 (6%) | 2 (9%) | 2 (4%) |
| Previously Healthy | 0 (0%) | 23 (33%) | 2 (33%) | 21 (32%) | 7 (30%) | 16 (33%) |
| Admitted | 0 (0%) | 15 (22%) | 1 (17%) | 14 (22%) | 8 (35%) | 7 (15%) |
| ICU admission | 0 (0%) | 3 (4%) | 1 (17%) | 2 (3%) | 3 (13%) | 0 (0%) |
| Need for respiratory support | 0 (0%) | 2 (3%) | 1 (17%) | 1 (2%) | 2 (9%) | 0 (0%) |
| Clinical Severity |  |  |  |  |  |  |
| Asymptomatic | 0 (0%) | 7 (10%) | 1 (17%) | 6 (9%) | 4 (17%) | 3 (6%) |
| Mild | 2 (100%) | 58 (85%) | 4 (67%) | 56 (88%) | 16 (70%) | 44 (94%) |
| Severe | 0 (0%) | 3 (4%) | 1 (17%) | 2 (3%) | 3 (13%) | 0 (0%) |

Supplementary Table 6**: Symptoms, grouped by SNP (excluding those with single type)**

|  |  | C241 | | C3037 | | A23403 | | C8782 | |
| --- | --- | --- | --- | --- | --- | --- | --- | --- | --- |
|  | Value | C | T | C | T | A | G | C | T |
| N | 71 | 22 | 49 | 15 | 56 | 12 | 59 | 69 | 2 |
| Asymptomatic | 8 (11%) | 5 (23%) | 3 (6%) | 4 (27%) | 4 (7%) | 4 (33%) | 4 (7%) | 8 (12%) | 0 (0%) |
| Fever or cough or shortness of breath | 57 (80%) | 16 (73%) | 41 (84%) | 10 (67%) | 47 (84%) | 7 (58%) | 50 (85%) | 55 (80%) | 2 (100%) |
| Fever | 38 (54%) | 10 (45%) | 28 (57%) | 6 (40%) | 32 (57%) | 3 (25%) | 35 (59%) | 37 (54%) | 1 (50%) |
| Cough | 41 (58%) | 11 (50%) | 30 (61%) | 7 (47%) | 34 (61%) | 6 (50%) | 35 (59%) | 39 (57%) | 2 (100%) |
| Shortness of Breath | 13 (18%) | 4 (18%) | 9 (18%) | 2 (13%) | 11 (20%) | 2 (17%) | 11 (19%) | 12 (17%) | 1 (50%) |
| Anosmia | 5 (7%) | 0 (0%) | 5 (10%) | 0 (0%) | 5 (9%) | 0 (0%) | 5 (8%) | 5 (7%) | 0 (0%) |
| Aguesia | 4 (6%) | 1 (5%) | 3 (6%) | 1 (7%) | 3 (5%) | 1 (8%) | 3 (5%) | 3 (4%) | 1 (50%) |
| Sore Throay | 13 (18%) | 2 (9%) | 11 (22%) | 0 (0%) | 13 (23%) | 0 (0%) | 13 (22%) | 13 (19%) | 0 (0%) |
| Chest pain | 4 (6%) | 1 (5%) | 3 (6%) | 1 (7%) | 3 (5%) | 1 (8%) | 3 (5%) | 3 (4%) | 1 (50%) |
| Myalgias | 12 (17%) | 1 (5%) | 11 (22%) | 1 (7%) | 11 (20%) | 0 (0%) | 12 (20%) | 12 (17%) | 0 (0%) |
| Chills | 5 (7%) | 1 (5%) | 4 (8%) | 1 (7%) | 4 (7%) | 1 (8%) | 4 (7%) | 4 (6%) | 1 (50%) |
| Headache | 23 (32%) | 5 (23%) | 18 (37%) | 3 (20%) | 20 (36%) | 2 (17%) | 21 (36%) | 22 (32%) | 1 (50%) |
| Fatigue | 7 (10%) | 1 (5%) | 6 (12%) | 0 (0%) | 7 (12%) | 0 (0%) | 7 (12%) | 7 (10%) | 0 (0%) |
| Gastrointestinal symptoms | 12 (17%) | 5 (23%) | 7 (14%) | 3 (20%) | 9 (16%) | 1 (8%) | 11 (19%) | 11 (16%) | 1 (50%) |

**Supplementary Table 6: Symptoms, grouped by SNP (excluding those with single type)**

|  |  | G25563 | | T28144 | | G28882 | |
| --- | --- | --- | --- | --- | --- | --- | --- |
|  | Total | G | T | C | T | A | G |
| N | 71 | 23 | 48 | 2 | 69 | 6 | 65 |
| Asymptomatic | 8 (11%) | 5 (22%) | 3 (6%) | 0 (0%) | 8 (12%) | 1 (17%) | 7 (11%) |
| Fever or cough or shortness of breath | 57 (80%) | 15 (65%) | 42 (88%) | 2 (100%) | 55 (80%) | 5 (83%) | 52 (80%) |
| Fever | 38 (54%) | 8 (35%) | 30 (62%) | 1 (50%) | 37 (54%) | 2 (33%) | 36 (55%) |
| Cough | 41 (58%) | 11 (48%) | 30 (62%) | 2 (100%) | 39 (57%) | 4 (67%) | 37 (57%) |
| Shortness of Breath | 13 (18%) | 3 (13%) | 10 (21%) | 1 (50%) | 12 (17%) | 1 (17%) | 12 (18%) |
| Anosmia | 5 (7%) | 0 (0%) | 5 (10%) | 0 (0%) | 5 (7%) | 0 (0%) | 5 (8%) |
| Aguesia | 4 (6%) | 1 (4%) | 3 (6%) | 1 (50%) | 3 (4%) | 0 (0%) | 4 (6%) |
| Sore Throat | 13 (18%) | 1 (4%) | 12 (25%) | 0 (0%) | 13 (19%) | 1 (17%) | 12 (18%) |
| Chest pain | 4 (6%) | 1 (4%) | 3 (6%) | 1 (50%) | 3 (4%) | 0 (0%) | 4 (6%) |
| Myalgias | 12 (17%) | 0 (0%) | 12 (25%) | 0 (0%) | 12 (17%) | 0 (0%) | 12 (18%) |
| Chills | 5 (7%) | 1 (4%) | 4 (8%) | 1 (50%) | 4 (6%) | 0 (0%) | 5 (8%) |
| Headache | 23 (32%) | 4 (17%) | 19 (40%) | 1 (50%) | 22 (32%) | 0 (0%) | 23 (35%) |
| Fatigue | 7 (10%) | 1 (4%) | 6 (12%) | 0 (0%) | 7 (10%) | 0 (0%) | 7 (11%) |
| Gastrointestinal symptoms | 12 (17%) | 3 (13%) | 9 (19%) | 1 (50%) | 11 (16%) | 1 (17%) | 11 (17%) |

Supplementary Table 7**. Comparison for Hospital admission at position 241, C versus T.**

|  | Odds Ratio | 95% Confidence Interval | P-value |
| --- | --- | --- | --- |
| Single Nucleotide Polymorphism |  |  |  |
| C241 | -ref- |  |  |
| T | .1365945 | 0.03-0.55 | 0.005 |
| Age in years | .9743804 | 0.87-1.09 | 0.653 |
| Sex |  |  |  |
| Female | -ref- |  |  |
| Male | 2.759766 | 0.66-11.55 | 0.165 |
| Race/Ethnicity |  |  |  |
| Non-Hispanic White | -ref- |  |  |
| Non-Hispanic Black | .5888435 | 0.09-3.94 | 0.585 |
| Other Race or Ethnicity | .3411012 | 0.03-3.99 | 0.391 |
| Insurance |  |  |  |
| Commercial Insurance | -ref- |  |  |
| Government or Public Insurance | 2.382647 | 0.34-16.54 | 0.380 |
| Other or Unknown | 1.040949 | 0.05-22.36 | 0.980 |

Model: SNP, age in years at time of testing, sex, race/ethnicity, and insurance payer

Supplementary Table 8**. Comparison for fospital admission at position 3037, C versus T.**

|  | Odds Ratio | 95% Confidence Interval | P-value |
| --- | --- | --- | --- |
| Single Nucleotide Polymorphism |  |  |  |
| C3037 | -ref- |  |  |
| T | 0.16 | 0.03-0.75 | 0.020 |
| Age in years | 0.98 | 0.87-1.09 | 0.670 |
| Sex |  |  |  |
| Female | -ref- |  |  |
| Male | 2.76 | 0.68-11.17 | 0.154 |
| Race/Ethnicity |  |  |  |
| Non-Hispanic White | -ref- |  |  |
| Non-Hispanic Black | 0.74 | 0.11-4.82 | 0.756 |
| Other Race or Ethnicity | 0.45 | 0.04-4.85 | 0.507 |
| Insurance |  |  |  |
| Commercial Insurance | -ref- |  |  |
| Government or Public Insurance | 3.21 | 0.53-19.51 | 0.206 |
| Other or Unknown | 0.92 | 0.04-19.46 | 0.955 |

Model: SNP, age in years at time of testing, sex, race/ethnicity, and insurance payer

Supplementary Table 9**. Comparison for Hospital admission at position 25563, G versus T.**

|  | Odds Ratio | 95% Confidence Interval | P-value |
| --- | --- | --- | --- |
| Single Nucleotide Polymorphism |  |  |  |
| G25563 | -ref- |  |  |
| T | 0.39 | 0.11-1.42 | 0.151 |
| Age in years | 0.97 | 0.87-1.07 | 0.527 |
| Sex |  |  |  |
| Female | -ref- |  |  |
| Male | 2.26 | 0.61-8.34 | 0.223 |
| Race/Ethnicity |  |  |  |
| Non-Hispanic White | -ref- |  |  |
| Non-Hispanic Black | 0.54 | 0.10-3.03 | 0.480 |
| Other Race or Ethnicity | 0.26 | 0.03-2.48 | 0.244 |
| Insurance |  |  |  |
| Commercial Insurance | -ref- |  |  |
| Government or Public Insurance | 3.24 | 0.56-18.75 | 0.190 |
| Other or Unknown | 2.38 | 0.15-38.60 | 0.541 |

Model: SNP, age in years at time of testing, sex, race/ethnicity, and insurance payer

Supplementary Table 10**. Comparison for Hospital admission at position 28882, G versus A.**

|  | Odds Ratio | 95% Confidence Interval | P-value |
| --- | --- | --- | --- |
| Single Nucleotide Polymorphism |  |  |  |
| G28882 | -ref- |  |  |
| G | 1.74 | 0.17-17.88 | 0.641 |
| Age in years | 0.94 | 0.85-1.05 | 0.258 |
| Sex |  |  |  |
| Female | -ref- |  |  |
| Male | 2.05 | 0.56-7.48 | 0.275 |
| Race/Ethnicity |  |  |  |
| Non-Hispanic White | -ref- |  |  |
| Non-Hispanic Black | 0.43 | 0.09-2.11 | 0.295 |
| Other Race or Ethnicity | 0.22 | 0.03-1.93 | 0.174 |
| Insurance |  |  |  |
| Commercial Insurance | -ref- |  |  |
| Government or Public Insurance | 3.56 | 0.69-18.47 | 0.130 |
| Other or Unknown | 2.57 | 0.16-40.07 | 0.500 |

Model: SNP, age in years at time of testing, sex, race/ethnicity, and insurance payer

Supplementary Table 11**. Comparison for Hospital admission at position 23403, A versus G.**

|  | Odds Ratio | 95% Confidence Interval | P-value |
| --- | --- | --- | --- |
| Single Nucleotide Polymorphism |  |  |  |
| A23403 | -ref- |  |  |
| G | .2280038 | 0.05-1.13 | 0.069 |
| Age in years | .962335 | 0.87-1.07 | 0.476 |
| Sex |  |  |  |
| Female | -ref- |  |  |
| Male | 2.68035 | 0.68-10.56 | 0.159 |
| Race/Ethnicity |  |  |  |
| Non-Hispanic White | -ref- |  |  |
| Non-Hispanic Black | .5535802 | 0.09-3.33 | 0.519 |
| Other Race or Ethnicity | .2766893 | 0.03-2.76 | 0.274 |
| Insurance |  |  |  |
| Commercial Insurance | -ref- |  |  |
| Government or Public Insurance | 4.201434 | 0.67-26.31 | 0.125 |
| Other or Unknown | 2.053693 | 0.12-35.23 | 0.620 |

Model: SNP, age in years at time of testing, sex, race/ethnicity, and insurance payer

Supplementary Table 12**. Comparison for lack of symptoms at position 23403, A versus G.**

|  | Odds Ratio | 95% Confidence Interval | P-value |
| --- | --- | --- | --- |
| Single Nucleotide Polymorphism |  |  |  |
| A23403 | -ref- |  |  |
| G | 0.11 | 0.01-0.92 | 0.042 |
| Age in years | 0.95 | 0.81-1.12 | 0.537 |
| Sex |  |  |  |
| Female | -ref- |  |  |
| Male | 0.15 | 0.01-1.87 | 0.140 |
| Race/Ethnicity |  |  |  |
| Non-Hispanic White | -ref- |  |  |
| Non-Hispanic Black | 0.67 | 0.04-10.41 | 0.778 |
| Other Race or Ethnicity | 4.43 | 0.24-80-94 | 0.316 |
| Insurance |  |  |  |
| Commercial Insurance | -ref- |  |  |
| Government or Public Insurance | 1.23 | 0.10-14.90 | 0.870 |
| Other or Unknown | 0.89 | 0.01-58.92 | 0.956 |

Model: SNP, age in years at time of testing, sex, race/ethnicity, and insurance payer
